# Pulmonary cavitation – an under-recognized late complication of severe COVID-19 lung disease

**DOI:** 10.1101/2020.08.15.20175869

**Authors:** Zaid Zoumot, Maria-Fernanda Bonilla, Ali S. Wahla, Irfan Shafiq, Mateen Uzbeck, Rania M. El-Lababidi, Fadi Hamed, Mohamed Abuzakouk, Mahmoud ElKaissi

## Abstract

**Background:** Radiological findings of the novel coronavirus disease 2019 (COVID-19) pulmonary disease have been well documented and range from scattered ground-glass infiltrates in milder cases to confluent ground-glass change, dense consolidation, and crazy paving in the critically ill, however, lung cavitation has not been described in these patients.

**Objectives:** To assess the incidence of pulmonary cavitation and describe its characteristics and evolution.

**Methods:** A retrospective review of all patients admitted to our institution with COVID-19 was undertaken and imaging reviewed to identify patients who developed pulmonary cavitation.

**Results:** Twelve out of 689 (1.7%) patients admitted to our institution with COVID-19 developed pulmonary cavitation, comprising 3.3% (n = 12/359) of those with COVID-19 pneumonia, and 11% (n = 12/110) of those admitted to the intensive care unit. We describe the imaging characteristics of the cavitation and present the clinical, pharmacological, laboratory, and microbiological parameters for these patients. In this cohort six patients have died while another remains critically ill and unlikely to survive.

**Conclusion:** Cavitary lung disease in patients with severe COVID-19 disease is not uncommon, and is associated with a high level of morbidity and mortality.

## INTRODUCTION

The novel coronavirus disease 2019 (COVID-19) pandemic has caused over 500,000 recorded deaths worldwide thus far[1]. Infection with the novel severe acute respiratory syndrome corona virus 2 (SARS-CoV-2) causes COVID-19 which can lead to pneumonia and severe acute respiratory syndrome. The typical abnormalities seen on computerized tomography (CT) of the chest in patients with COVID-19 lung disease have been well described,[2–5] with a comprehensive review and meta-analysis [6] of 55 studies finding peripheral ground glass opacities in most, consolidation in 44% (95% CI: 1–71%), air bronchograms in 43% (95% CI: 8–80%), linear opacities in 41% (95% CI: 7–65%), crazy-paving pattern in 24% (95% CI: 3–92%) and interlobular septal thickening in 23% (95% CI: 1–80%) of the CT scans reviewed. Notable in this meta-analysis and other studies is the absence of cavitation. Similarly, another meta-analysis of 15 studies including 2451 patients did not report any cavitation, but commented on the development of traction bronchiectasis, consolidation, lymphadenopathy and pleural effusions at late stages of severe disease[7]. Here, we present a series of 12 cases of SARS-CoV2 infection that had developed cavitary lung disease while admitted to our tertiary care hospital for treatment of severe COVID-19.

### MATERIALS AND METHODS

A registry of all patients admitted to our institution with COVID-19, and approved by the institution’s Research Ethics Committee, was retrospectively reviewed. In total 689 patients were admitted between February 23^rd^ and July 3^rd^ 2020, of whom 330 had asymptomatic or mild disease, 359 had evidence of pneumonia and 110 were admitted to Intensive Care for treatment of respiratory failure. One hundred and seventy eight patients had CT scans of the chest and all patients who had cavitary lung disease on CT scan were identified. Baseline demographics, comorbidities, symptoms, ventilatory and other Intensive care parameters, microbiology, medications received and hospital outcomes were reviewed. Data was extracted by one of three members of the research team, and verified by a second researcher. All CT scans were reviewed by a single thoracic radiologist and imaging characteristics were noted.

In the absence of approved pharmacologic therapy for COVID-19, the Department of Infectious Diseases and the antimicrobial stewardship program at our institution rapidly developed institutional practice guidelines for COVID-19 based on available in-vitro and clinical studies to aid clinicians with treatment decisions. These guidelines were frequently updated based on emerging data. In the early stages of the pandemic asymptomatic and symptomatic patients were treated with a combination of two or three medications that included hydroxychloroquine, lopinavir-ritonavir and favipiravir, for a duration of 5-10 days. Tocilizumab was recommended for those with evolving cytokine release storm (CRS) based on criteria that encompassed clinical, radiological and laboratory parameters. One dose was initially given and a second dose was optional based on the clinical evolution of the patient. Empiric antibacterial and antifungal treatment was not routinely administered unless there was clinical suspicion for secondary pneumonia. Amoxicillin-clavulanate and piperacillin-tazobactam were the most common antibiotics used for COVID-19 patients.

### RESULTS

Twelve patients with COVID-19 disease developed lung cavitation, comprising 1.7% (n = 12/689) of all admissions, 3.3% (n = 12/359) of those with COVID-19 pneumonia, and 11% (n = 12/110) of those admitted to the ICU. Table 1 describes their baseline characteristics, clinical variables and outcomes. Median (range) age was 47 (37 to 67) years, 50% (n = 6/12) had diabetes mellitus, 42% (n = 5/12) were hypertensive and one had chronic lung disease in the form of chronic obstructive pulmonary disease (COPD). None had received prior immunosuppression. All 12 were males, required invasive mechanical ventilation, and the median (range) time from symptom onset to invasive mechanical ventilation was 8 (3-15) days. Three were admitted directly via our institution’s Emergency Department and intubated within 24 hours, whilst the other nine patients were transferred from other hospitals within 72 hours of intubation. At the time of this data review six of the 12 patients had died, another remains critically ill and unlikely to survive, one is still admitted having been recently decannulated off extra-corporeal membrane oxygenation (ECMO) and had his tracheostomy decannulated, and four have been discharged home.

**Table 1:** Patient characteristics and outcomes of those who developed pulmonary cavitation.

|  | Patient 1 | Patient 2 | Patient 3 | Patient 4 | Patient 5 | Patient 6 | Patient 7 | Patient 8 | Patient 9 | Patient 10 | Patient 11 | Patient 12 |
| --- | --- | --- | --- | --- | --- | --- | --- | --- | --- | --- | --- | --- |
| Age | 54 | 44 | 67 | 49 | 60 | 51 | 42 | 38 | 42 | 50 | 36 | 42 |
| BMI | 30.9 | 30.4 | 31.3 | 29.2 | 25.1 | 29.4 | 26.3 | 21.5 | 30.1 | 19.8 | 24 | 27.7 |
| Comorbidities | DM, HT | HT | DM | DM | COPD | HT | HT |  | DM, HT | DM | DM | DM |
| On admission |  |  |  |  |  |  |  |  |  |  |  |  |
| Time from symptom onset to intubation (days) | 6 | 12 | 9 | 7 | 5 | 5 | 8 | 15 | 3 | 9 | 10 | 8 |
| P/F ratio | 48.8 | 41.6 | 49.7 | 33.0 | 46.1 | 47.0 | 65.3 | 41.8 | 70.5 | 100.1 | 109.4 | 104.5 |
| SOFA score (points) | 24 | 21 | 24 | 25 | 23 | 23 | 16 | 21 | 25 | 23 | 18 | 17 |
| Neutropenia |  |  |  |  |  |  |  |  |  |  | Y |  |
| Leucocytes (x10<sup>9</sup>/L) | 0.49 | 1.35 | 0.3 | 2.78 | 0.6 | 1.1 | 1.66 | 1.03 | 0.46 | 2.91 | 1.62 | 2.63 |
| During admission |  |  |  |  |  |  |  |  |  |  |  |  |
| NM blockade | Y | Y | Y | Y | Y | Y | Y | Y | Y |  |  | Y |
| VTE |  | DVT and PE |  |  |  | PE | DVT |  |  | DVT |  |  |
| CVA | Y |  |  |  |  |  |  |  |  | Y |  |  |
| CRRT/IHD |  | Y | Y | Y | Y | Y |  |  | Y |  |  | Y |
| ECMO |  |  |  |  |  |  | Y | Y | Y |  |  |  |
| Proned | Y | Y | Y | Y | Y | Y |  | Y | Y | Y |  |  |
| Tracheostomy | Y |  | Y | Y | Y | Y | Y | Y | Y |  |  |  |
| Days of systemic CS | 17 | 15 | 19 | 24 | 11 | 14 | 14 | 25 | 18 | 9 | 6 | 5 |
| +ve fungal cultures or serology |  | Y | Y | Y |  | Y |  |  | Y |  |  |  |
| Treated for fungal infection | Y | Y | Y | Y |  | Y | Y |  | Y |  |  | Y |
| Duration of hospital stay (days) | 37 | 25 | 53 | 33 | 53 | 74 | 56 | 57 | 57 | 56 | 47 | 40 |
| Outcome |  |  |  |  |  |  |  |  |  |  |  |  |
|  | Deceased | Deceased | Deceased | Deceased | Deceased | Deceased | Unlikely to survive | Likely to survive | Discharged home | Discharged home | Discharged home | Discharged home |

BMI= Body Mass Index, P/F ratio = PaO<sub>2</sub>/FiO<sub>2</sub>, SOFA = Sequential Organ Failure Assessment, NM = neuromuscular, Y=yes, VTE = venous thromboembolism, DVT= deep vein thrombosis, PE=pulmonary embolus, CVA = cerebrovascular accident, CRRT = continuous renal replacement therapy, IHD = intermittent hemodialysis, ECMO = extracorporeal membrane oxygenation, CS= corticosteroids

Most patients had completed a course of Hydroxychloroquine (83 % n = 10/12) and antivirals (favipilavir or lopinavir/ritonavir) (92%, n = 11/12) in the early stages of their illness. All 12 patients received Tocilizumab as clinical evidence of a CRS became apparent accompanied by deteriorating respiratory status and progressively worsening disease on imaging. Overall, this represented 9% (n=12/133) of patients with COVID-19 who received tocilizumab in our institution.

#### Imaging characteristics

Upon admission all patients had baseline chest X-rays which did not reveal any evidence of cavitary disease. The 12 patients had the recognized range of imaging features expected in severe COVID-19 lung disease. All had bilateral disease with ground glass opacities predominantly in the peripheries some with centrally located opacities, most had consolidation and air bronchograms, and roughly half had crazy paving pattern and interlobular septal thickening. In 10 patients all five lobes were affected, and in two patients the disease spared two lobes. The median (range) number of days between symptom onset and the first CT demonstrating cavitation was 36 (21 to 54) days, and between intubation and the first CT demonstrating cavitation 28 (13 to 49) days. Table 2 describes the cavities in detail. In short, five of the 12 patients had solitary cavities with size ranging between 30 mm to 100 mm in diameter. All patients with more than one cavity had bilateral cavitation. The appearances and morphology of the cavities amongst the group were similar with to that of pulmonary abscesses with thick but smooth walls containing internal debris and air fluid levels. Pulmonary infarcts were excluded as the CT studies were performed with IV contrast excluding pulmonary emboli. All five lobes contained cavities in a similar proportion, with a predilection for the costophrenic angles and the apices.

**Table 2.** Characteristics of the pulmonary cavities and related events.

|  | Patient 1 | Patient 2 | Patient 3 | Patient 4 | Patient 5 | Patient 6 | Patient 7 | Patient 8 | Patient 9 | Patient 10 | Patient 11 | Patient 12 |
| --- | --- | --- | --- | --- | --- | --- | --- | --- | --- | --- | --- | --- |
| No. of Cavities (n) | 1 | 8 | 8 | 3 | 1 | 1 | 2 | 1 | 9 | 3 | 1 | 5 |
| Largest Cavity (mm) | 50 | 85 | 60 | 30 | 30 | 30 | 70 | 100 | 54 | 52 | 50 | 40 |
| Bilateral |  | Y | Y | Y |  |  | Y |  | Y | Y |  | Y |
| Location of Cavities |  |  |  |  |  |  |  |  |  |  |  |  |
| No. lobes with cavities (n) | 1 | 5 | 5 | 3 | 1 | 4 | 2 | 1 | 4 | 3 | 1 | 3 |
| RUL |  | Y | Y | Y |  |  |  | Y | Y | Y | Y |  |
| RML |  | Y | Y |  | Y | Y |  |  | Y |  |  |  |
| RLL |  | Y | Y |  |  | Y | Y |  | Y | Y |  | Y |
| LUL |  | Y | Y | Y |  | Y | Y |  | Y |  |  | Y |
| LLL | Y | Y | Y | Y |  | Y |  |  |  | Y |  | Y |
| Clinical Events |  |  |  |  |  |  |  |  |  |  |  |  |
| Developed Pneumothorax |  |  |  | Y |  |  |  | Y | Y |  |  | Y |
| Developed Hemoptysis |  |  | Y |  |  |  | Y |  | Y |  |  | Y |
| Treated for Invasive fungal infection | Y | Y | Y | Y |  | Y | Y |  | Y |  |  | Y |
| Bacterial organisms in Sputum/BAL | *K. pneumoniae* | *ECC* | *MRSA* |  | *S. maltophilia*,<br>*C. koseri* | *MSSA*,<br>*S. maltophilia* | *S. marcescens*,<br>*S. maltophilia*,<br>*ECC* | *K. pneumoniae* | *Acinetobacter*,<br>*MRSA* | *ESBL K. pneumoniae*,<br>*MRSA* | *MRSA* | *K. pneumoniae* |

Y= Yes, RUL = Right upper lobe, RML = Right middle lobe, RLL = Right lower lobe, LUL = Left upper lobe, LLL = Left lower lobe, BAL = Bronchoalveolar Lavage, MRSA = Methicillin-resistant Staphylococcus aureus, MSSA = Methicillin-sensitive Staphylococcus aureus, K. pneumoniae = Klebsiella pneumonia, ECC = Enterobacter cloacae complex, S. maltophilia = Stenotrophomonas maltophilia, S. marcescens = Serratia marcescens, ESBL = Extended spectrum beta-lactamase

### DISCUSSION

The development of pulmonary cavitation in patients with severe COVID-19 lung disease treated in our institution’s ICU was not a rare event (11%, n = 12/110). This subgroup of patients had very severe infection with acute respiratory distress syndrome (ARDS) and required a prolonged ICU stay. Median (range) Sequential Organ Failure Assessment (SOFA) score was 23 (range 16-24) on admission, and all patients were leucopenic. Seven of 12 required renal replacement therapy, four developed venous thromboembolism, three required ECMO with two surviving to successful decannulation, and two had thromboembolic cerebrovascular events.

Cavities tended to form in areas of the lung where ground glass opacities where seen in early stages, morphing into more dense consolidation, later developing necrosis and ultimately cavitating. This is demonstrated for patients 8 and 9 in Figures 1 and 2, respectively. We are unable to speculate as to whether bacterial infection and/or invasive fungal coinfection may have contributed to the development of the cavities, or if the infections were opportunistic. However it is uncommon for viral pneumonias,[8] including those due to the other human coronaviruses SARS-CoV[9] and MERS-CoV,[10] to cause pulmonary cavitation even in severe and advanced viral infection. Furthermore, four of twelve patients who had developed pulmonary cavitation (including two of the survivors) had no microbiological, serological, clinical or distinct radiological characteristics of invasive fungal infection and did not receive treatment for this.

**Fig. 1.**
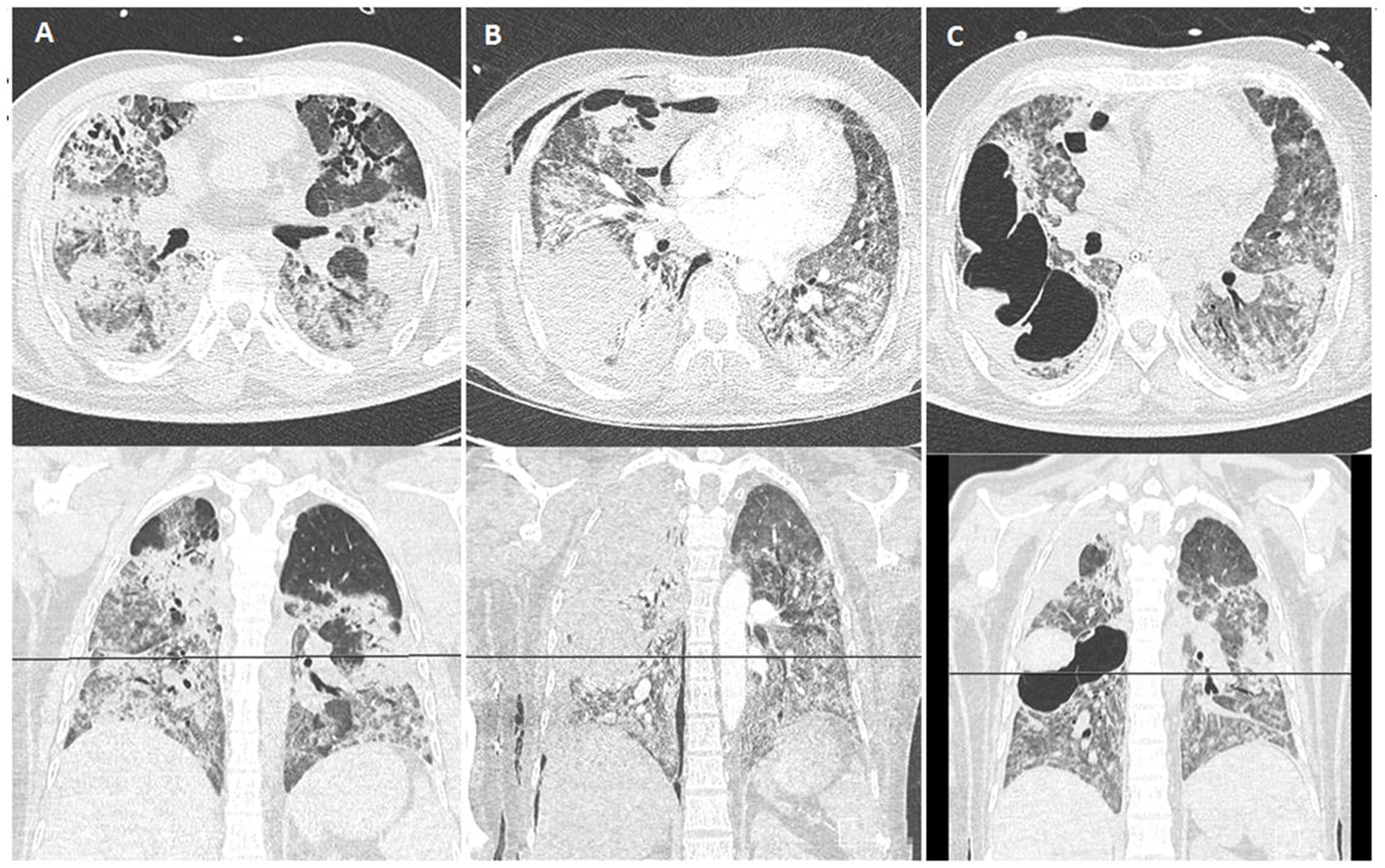
Axial and coronal CT images at form patient 8 on (A) 5^th^ May 2020, (B) 26^th^ May 2020, and (C) 6^th^ June 2020.

**Fig. 2.**
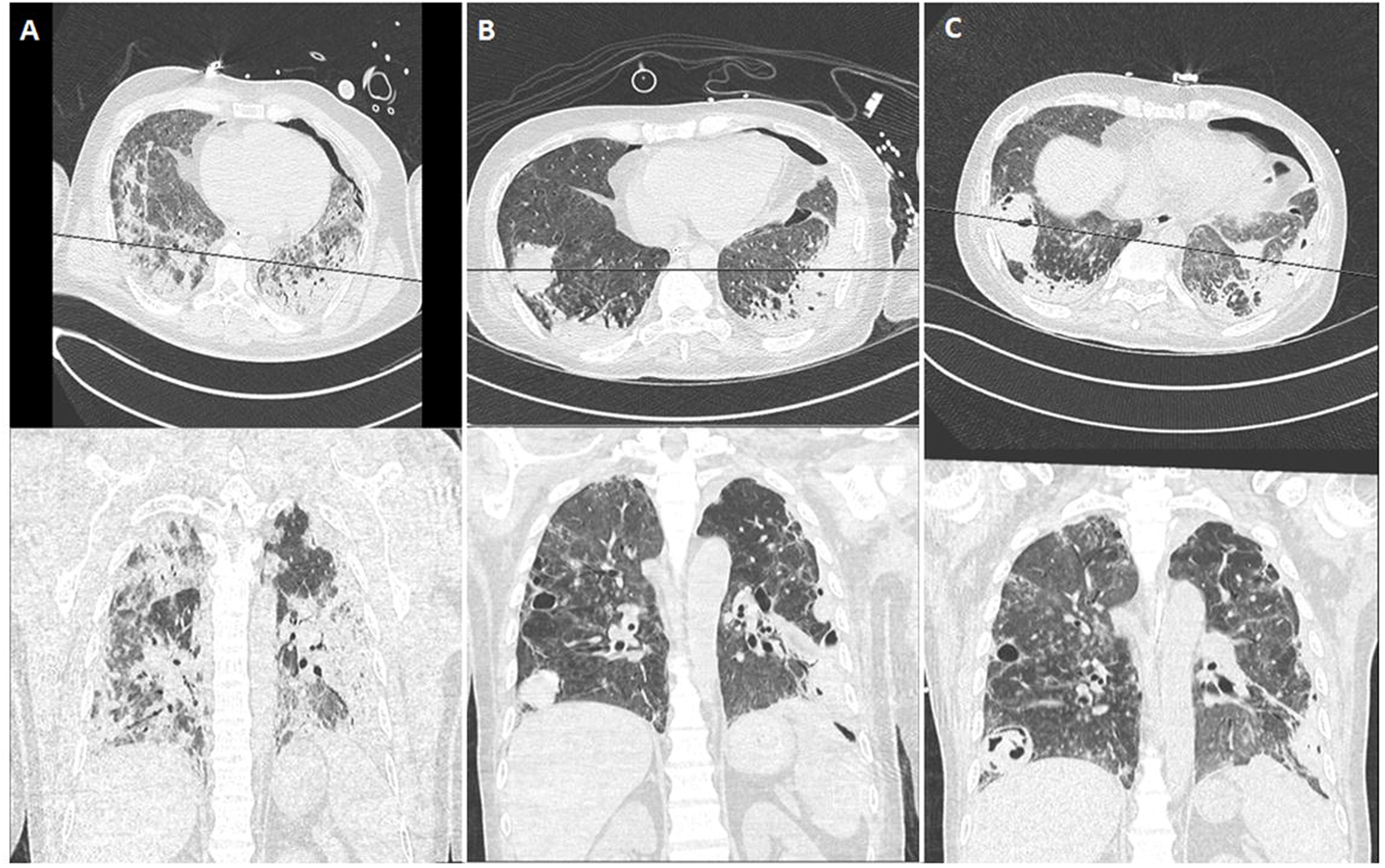
Axial and coronal CT images at form patient 9 on (A) 14^th^ May 2020, (B) 8^th^ June 2020, and (C) 18^th^ June 2020.

Four patients developed hemoptysis and all had features of suspected invasive aspergillosis. Hemoptysis appeared to have occurred irrespective of cavity size. Similarly, secondary pneumothorax also occurred in patients with both larger and smaller cavities.

All patients in this series received tocilizumab, a recombinant humanized monoclonal antibody directed against both the soluble and membrane-bound forms of the interleukin-6 (IL-6) receptor, in the early stages of a CRS. Tocilizumab is currently approved by the US Food and Drug Administration (FDA) for the treatment of severe rheumatoid arthritis, systemic juvenile idiopathic arthritis, giant cell arteritis, and life-threatening CRS induced by chimeric antigen receptor T cell therapy[11]. Recently it has been associated with improved survival in patients with severe COVID-19 pneumonia with evidence of CRS.[12,13] In general tocilizumab is well tolerated but can induce neutropenia, and an increased risk of developing infections has been reported.[14,15] Furthermore, it may predispose to a delay in detecting active infection because of the masking effect of a suppressed C reactive protein (CRP) response. Interestingly, however, only one of 12 patients in our cohort developed neutropenia during their ICU stay.

All patients in this series also received systemic glucocorticoids, which may have survival benefit in COVID-19[16], but also suppress the immune system by impairing innate immunity. In the treatment of patients reported here systemic steroids were administered as part of our ICU protocol for septic shock, based on the Society of Critical Care Medicine guidelines[17] and not directly for the treatment of their COVID-19.

The high level of morbidity and mortality in this small case series highlights that cavity formation probably sits at the severe/end-stage of the radiological COVID-19 spectrum. It is unclear what the natural history of these cavities will be in survivors. This will be informed by future follow-up interval imaging but it is reasonable to assume that though there may be some of regression in the size of the cavities, there will be an increased risk of pneumothorax, hemoptysis, colonization with bacteria including non-tuberculous mycobacteria, fungi and the development of mycetomas in the future.

## CONCLUSION

This study highlights that pulmonary cavitation in patients with severe COVID-19 lung disease can occur, is associated with secondary complications of hemoptysis and pneumothorax, and confers a poor prognosis. Early cross sectional imaging should be considered if there is suspicion of cavitation on plain radiographs, and a more aggressive investigation and treatment of possible invasive fungal infection undertaken. Further studies are needed to determine whether treatment with tocilizumab, systemic glucocorticoids or a combination of both may increase the risk of developing pulmonary cavitation in patients with COVID-19.

## Data Availability

All data in the manuscript are available upon request.

## Statement of Ethics

Our institution’s Research Ethics Committee approved this retrospective review of an authorized registry of all patients admitted with COVID-19.

## Conflict of Interest Statement

The authors have no conflicts of interest to declare.

## Funding Sources

There was no funding for this study.

## Author Contributions

Z.Z. conceived this study, collated data and wrote the first draft of the manuscript and all authors reviewed and contributed to the writing of subsequent drafts, and approved the final version. M-F.B., A.S.W., I.S., M.U., and R.M.E assisted in data collection and M.E. performed the imaging review, data collection and analysis.

## Notes

### Competing Interest Statement

The authors have declared no competing interest.

### Author Declarations

Cleveland Clinic Abu Dhabi Research Ethics Committee.

